# Health conditions associated with sexual assault in a large hospital population

**DOI:** 10.1101/2022.04.04.22273398

**Authors:** Allison M. Lake, Slavina B. Goleva, Lauren R. Samuels, Laura M. Carpenter, Lea K. Davis

## Abstract

**Objective:** To develop a clinical informatics approach to identify patients with a history of sexual assault and to characterize the clinical risk factors and comorbidities of this population in a sex-stratified manner.

**Methods:** We developed and applied a keyword-based approach to clinical notes to identify patients with a history of sexual assault in the Vanderbilt University Medical Center (VUMC) electronic health record from 1989 to 2021. Using a phenome-wide association study (PheWAS), we then examined diagnoses that co-occurred with evidence of sexual assault. We also examined whether sex assigned at birth modified any of these associations.

**Results:** Our keyword-based algorithm achieved a positive predictive value of 90.4%, as confirmed by manual patient chart review. Out of 1,703 diagnoses tested across all subgroup analyses, we identified a total of 465 associated with sexual assault, many of which have been previously observed in the literature. Interaction analysis revealed 55 sex-differential phenotypic associations.

**Conclusions:** In a large hospital setting, disclosures of sexual assault were associated with increased rates of hundreds of health conditions.

## Introduction

Sexual assault, which includes rape, forced penetration of another person, sexual coercion, and unwanted sexual contact, is a major public health and human rights concern, estimated to affect 44% of women and 25% of men in the United States during their lifetimes^1^. Rape alone has an astonishing lifetime prevalence of 20% in women^1^. Sexual assault has immediate and wide-ranging health consequences, including direct physical injuries and adverse effects on reproductive and mental health^2^. Sexual assault is also associated with debilitating long-term health outcomes. Survivors of sexual assault exhibit increased rates of psychiatric conditions including post-traumatic stress disorder (PTSD), anxiety, depression, sleep disorders, eating disorders, and suicide attempts^3^. Furthermore, sexual assault is associated with lifetime diagnoses of multiple functional and somatic disorders, including gastrointestinal symptoms, chronic pain, and functional seizures^4^.

The ability to identify patients with a history of sexual assault is crucial for understanding health consequences related to sexual assault. Large studies are needed to understand the unique healthcare needs of this population. However, the challenges inherent in sexual assault research make this a daunting task. The highly sensitive and often stigmatized nature of sexual assault can leave many patients and research participants unwilling to disclose these experiences. Stigmatization and other cultural factors may also lead survivors to reject the classification of their experiences as sexual assault^5^ (cited in^6^). Furthermore, research on male survivors is relatively limited, likely in part due to differential reporting of sexual assault between men and women^7^. Given the sex differences widely observed for common mental health conditions, it is plausible that the effects of sexual assault on mental health differ between men and women. Despite the importance of this line of research, sex differences in sexual assault-related health effects are understudied, and few robust sex differences have been identified^3,4,7^.

Research utilizing electronic health records (EHRs) has the potential to overcome some limitations of traditional epidemiological approaches. However, few EHR-based studies of sexual assault and associated health sequelae exist^8,9^. In the present study, we utilized both billing codes and key phrases to identify patients with a history of sexual assault in a large hospital setting. Our novel key phrase approach, which mines clinical note text for matches to certain relevant phrases, identified over 14,000 patients who have experienced sexual assault out of over 3 million patients, a twofold increase over the 6,998 patients identified using billing codes alone. We then characterized the medical phenome of a subset of this patient population and identified hundreds of associated clinical phenotypes, including many related to psychiatric conditions. Finally, we demonstrated the sex-differentiated nature of dozens of clinical associations, addressing a key gap in sexual assault research.

## Methods

### Sample and data description

This study included 3,376,424 de-identified patients who received care at VUMC, an academic medical center in Nashville, TN, between the years 1989 and 2021. Demographic information, ICD-9 and ICD-10 codes, and clinical notes were obtained from the medical record for use in subsequent analyses. No age restrictions were applied. For all association analyses, we retained only individuals meeting “medical home” criteria as described^10^ to limit the study population to patients receiving regular care at VUMC and reduce nonrandom missingness between cases and controls. This was accomplished by restricting our study population to individuals with at least 5 ICD-9 or ICD-10 codes, of any type, on different days over a period of at least 3 years. We further limited the study population to individuals with recorded male or female sex assigned at birth, and with at least one recorded BMI. This study was reviewed and approved by the VUMC institutional review board (IRB #212285) and exempted from informed consent requirements as non-human subjects research using deidentified medical data.

### Sexual assault case/control algorithms

Although ICD codes do exist for rape, sexual assault, and sexual abuse, these are typically reserved for care immediately after an assault and are infrequently used to indicate a history of sexual abuse or assault. Across the entire cohort, only 6,998 patient charts included ICD codes for sexual assault (0.21%), likely an extreme underestimate given sexual assault prevalence estimates^1^. To improve detection of patients with a history of sexual assault, we developed an algorithm to search the unstructured free text from clinical notes associated with patient charts for matches to specific phrases. These clinical notes consisted of admission notes, ancillary reports, discharge summaries, emergency department notes, inpatient notes, notes not otherwise classified, nursing reports, outpatient notes, pathology reports, and problem lists.

In developing the algorithm, we first identified charts containing relevant ICD codes and/or matches to an initial set of keywords with obvious relevance (e.g., “rape” and “sexual assault”). We then performed an exploratory chart review and identified an expanded set of relevant keywords and exclusion phrases. This subsequent “phase 1” algorithm identified individuals meeting criteria for disclosure of sexual assault using the chart-review-derived set of key phrases as well as relevant ICD codes (**Supplemental Table 1**). We then performed a comprehensive manual review of 25 charts meeting these criteria to further refine our keyword search terms accordingly. This included identifying commonly used phrasing to indicate the presence of a history of sexual assault (i.e., inclusion) or the absence of a history of sexual assault (i.e., exclusion).

Our final “phase 2” keyword-based algorithm consisted of 18 inclusion phrases (e.g., “history of sexual assault”, “hx of rape”) and 12 exclusion phrases (i.e., “denies history of sexual abuse”, “no hx of sexual assault”) to identify patients who disclosed sexual assault (**Supplemental Table 1**). To allow comparisons between keyword-based and ICD-based approaches, ICD codes were not used in this final keyword-based algorithm. A patient was considered a case if their chart contained a match to at least one inclusion phrase and contained no matches to any exclusion phrases. Otherwise, the patient was considered a control.

To benchmark our keyword-based algorithm, we performed additional analyses using only ICD codes to identify patients with positive and negative histories of sexual assault. In the ICD-code-based approach, charts were defined as cases if they contained at least 1 of 25 ICD-9 and ICD-10 codes pertaining to sexual assault (**Supplemental Table 2**), and as controls otherwise. We did not exclude individuals meeting both ICD-based and keyword-based criteria.

### Chart review

To evaluate our final “phase 2” keyword-based algorithm, AML conducted a manual review of 52 randomly selected charts identified by the sexual assault algorithm and calculated the positive predictive value of the algorithm. The chart reviews were conducted within a subset of patients who are also enrolled in the VUMC biobank, BioVU. These patients tend to have longer health records (median 9.99 years among biobank participants compared to median of 1.15 years for the entire VUMC-EHR) and thus present an optimal setting for validating a phenotyping algorithm that relies on the presence of key phrases in clinical notes. Charts were identified as true positives if they clearly contained a description of the patient’s reported history of sexual assault.

### Phecode-based analysis

The presence or absence of a history of sexual assault was used as the primary independent variable in a PheWAS. The medical phenome was categorized by phecodes—single codes representing clinical phenotypes—that were generated by aggregating related billing codes into hierarchical code families^11^. Using the PheWAS package^12^ (version 0.99) in R^13^, we mapped ICD-9 and ICD-10 codes to 1,817 phecodes using Phecode Map version 1.2 (phewascatalog.org)^14,15^. We conducted a sex-combined PheWAS and sex-stratified (i.e., separately in males and females) PheWAS. For each phenotype tested in the sex-combined analysis, we required a minimum of 200 records, with at least 100 records for both males and females. Accordingly, for the sex-stratified analyses, we required a minimum of 100 records per phecode. For each phecode analyzed (n=1482, 1658, and 1527 for sex-combined, female-only, and male-only, respectively), we fitted a multiple logistic regression model with sexual assault history (binary: positive or negative) as the primary independent variable and EHR-reported race, ethnicity, median age of all visits in an individual’s medical record, median body mass index (BMI) of all visits in an individual’s medical record, and the log-transformed mean number of ICD codes per day in the record as covariates. Sex-combined analyses additionally included sex as a covariate.

For the 354 phenotypes that were significantly associated with sexual assault in at least one sex in the sex-stratified analyses and that had at least 100 records for each sex, we conducted interaction analyses to formally examine the moderating effect of sex on phenotypic associations with sexual assault. These analyses used models identical to those used in the sex-combined analyses, with the addition of a sex-by-sexual-assault interaction term.

For each association analysis, we applied a Bonferroni correction (p= 0.05/total number of phecodes tested) to correct for the number of phecodes tested, using a threshold of corrected p<0.05 for an association to be considered phenome-wide-significant. Although Bonferroni correction is likely to be overly conservative given the correlation inherent in related diagnostic codes, the large sample size counteracts this loss of power.

## Results

### Sexual assault prevalence in the electronic health record

Across our entire cohort (3,376,424 patients), the keyword-based approach identified 14,496 individuals (0.43%) with documentation of sexual assault, whereas only 6,998 cases (0.21%) (with an overlap of 2,928 individuals) received ICD codes for sexual assault. After applying medical-home criteria to our cohort (see *Methods*), we removed 11 individuals with unknown sex and 154,804 individuals with missing BMI, resulting in a total sample of 833,185 individuals. Among these individuals, 9,333 were designated as cases by the keyword-based algorithm (0.94%, **Table 1**), and 4,422 by the ICD-code-based algorithm (0.45%, **Supplemental Table 3**), with an overlap of 2,014 individuals. Patients with keyword- or ICD-based documentation of sexual assault were predominantly female, and younger on average than the remainder of the medical-home population (**Table 1, Supplemental Table 3**).

**Table 1:** Demographics of patients classified as sexual assault cases or controls by our keyword-based phenotyping algorithm. To assess case-control differences in demographic variables, two-sided t-tests were performed for continuous variables (record-median age, record-median BMI, log(mean records per day)), and chi-square tests were performed for binary variables (sex, white/non-Hispanic). Log(mean records per day) is a measure of the density of diagnostic codes across an individual’s health record.

|  | Case<br>(N=9333) | Control<br>(N=823852) | Test<br>statistic | P-value |
| --- | --- | --- | --- | --- |
| **Sex** |  |  |  |  |
| Female | 7610 (81.5%) | 467617 (56.8%) | $\chi^2=2311.26$ | <0.001 |
| Male | 1723 (18.5%) | 356235 (43.2%) |  |  |
| **Record-median age** |  |  |  |  |
| Median [Min, Max] | 28.0 [0, 86.0] | 37.5 [0, 89.0] | t=-39.47 | <0.001 |
| **Record-median BMI** |  |  |  |  |
| Median [Min, Max] | 26.4 [11.3, 59.2] | 25.8 [10.0, 60.0] | t=18.04 | <0.001 |
| **White and non-Hispanic** |  |  |  |  |
| No | 2436 (26.1%) | 207663 (25.2%) | $\chi^2=3.87$ | 0.0492 |
| Yes | 6897 (73.9%) | 616189 (74.8%) |  |  |
| **Log(mean records per day)** |  |  |  |  |
| Median [Min, Max] | -3.44 [-7.16, 0.699] | -4.09 [-7.68, 1.37] | t=53.07 | <0.001 |

### Sexual assault case-control algorithm performance

Of the 52 patient charts identified as cases by the sexual assault case-control algorithm and randomly selected to assess the positive predictive value (PPV) of the algorithm, 47 were found to be true positives (PPV=90.4%, 95% confidence interval [CI] 78.2%-96.4%). In the 5/52 charts identified as false positives, inclusion phrases were present either as negation of sexual assault history (e.g., “denies a h/o sexual abuse”) or in reference to someone other than the patient (e.g., “[sibling] was sexually abused”).

### History of sexual assault is associated with hundreds of clinical phenotypes

Our sex-combined PheWAS identified 386 out of 1,482 medical and psychiatric conditions as significantly overrepresented (Bonferroni-adjusted α=3.34e-05) among patients disclosing sexual assault. Of the 50 associations with lowest p-values, 40 were psychiatric disorders, including schizophrenia (OR=86.36, 95% CI=76.31, 97.75), suicide or self-inflicted injury (OR=86.33, 95% CI=74.82, 99.6), depression (OR=27.5, 95% CI=25.57, 29.57), and PTSD (OR=122.81, 95% CI=113.47, 132.91, **Figure 1, Supplemental Table 4**). These top associations also included phecodes relating to personality disorders: “personality disorders” (OR=107.35, 95% CI=94.51, 121.92) and “antisocial/borderline personality disorder” (OR=129.17, 95% CI=111.73, 149.32). In total, 64 of the 71 tested phecodes pertaining to psychiatric disorders were significantly associated with sexual assault.

**Figure 1:**
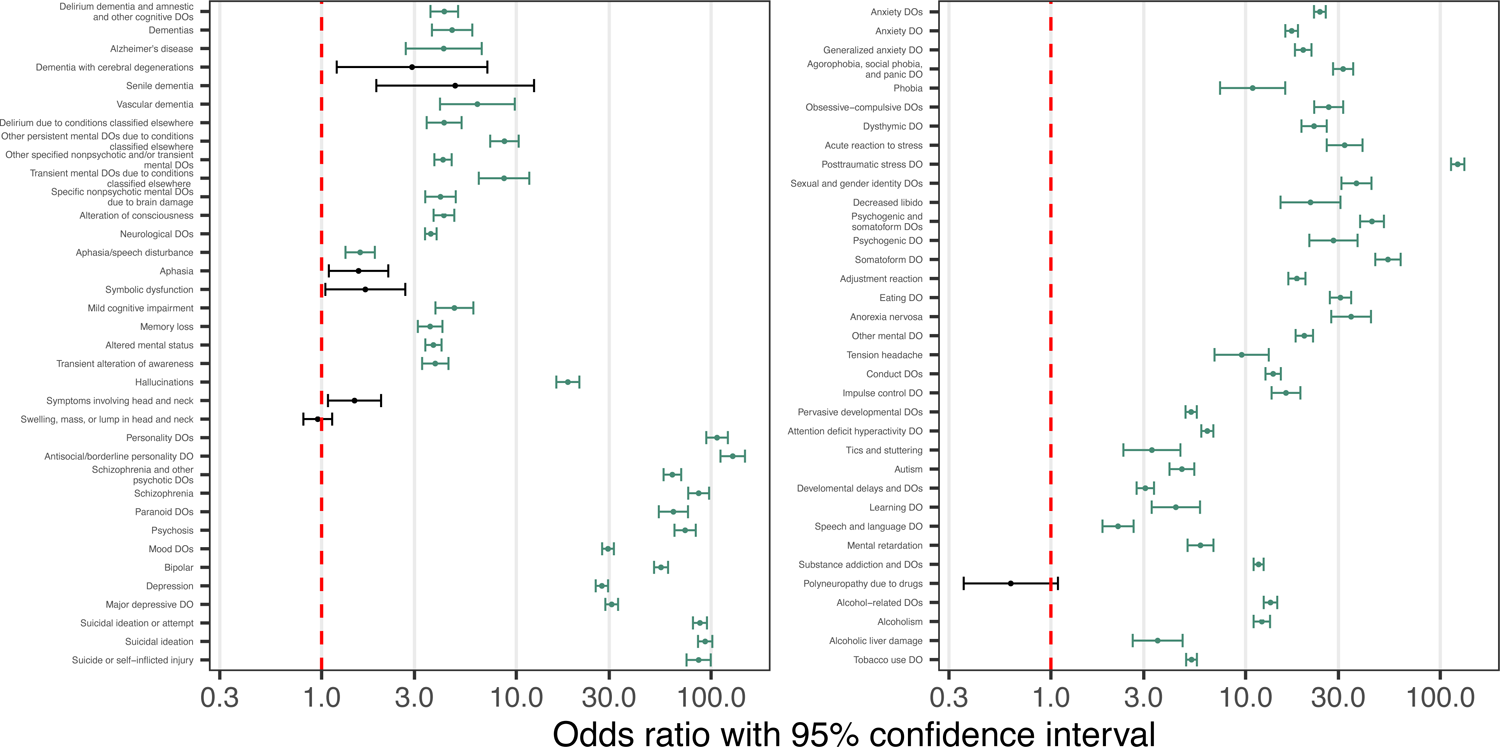
Psychiatric phenotypic associations with sexual assault. Log10-scale forest plots displaying odds ratios with 95% confidence intervals for adjusted associations between sexual assault and all tested psychiatric phecodes in sex-combined analyses. Associations achieving statistical significance are colored green. Related phecodes are grouped together (for example, the “Antisocial/borderline personality DO” phecode [301.2] is related to its parent phecode “Personality DOs” [301]). Red dashed line indicates an OR of 1. (Note: DO=disorder).

The 386 statistically significant associations also included many non-psychiatric health conditions. Some of these associations plausibly represent immediate physical consequences of sexual assault, including urinary tract infection (OR=1.79, 95% CI=1.68, 1.91), sexually transmitted infection (OR=4.6, 95% CI=3.85, 5.5), and contusion (OR=2.19, 95% CI=1.9, 2.53), **Figure 2, Supplemental Table 4**). Many of the non-psychiatric clinical associations with the largest effect sizes pertained to poisonings, including poisoning by psychotropic agents (OR=18.13, 95% CI=15.55, 21.13) and poisoning by analgesics, antipyretics, or antirheumatics (OR=5.35, 95% CI=4.69, 6.09).

**Figure 2:**
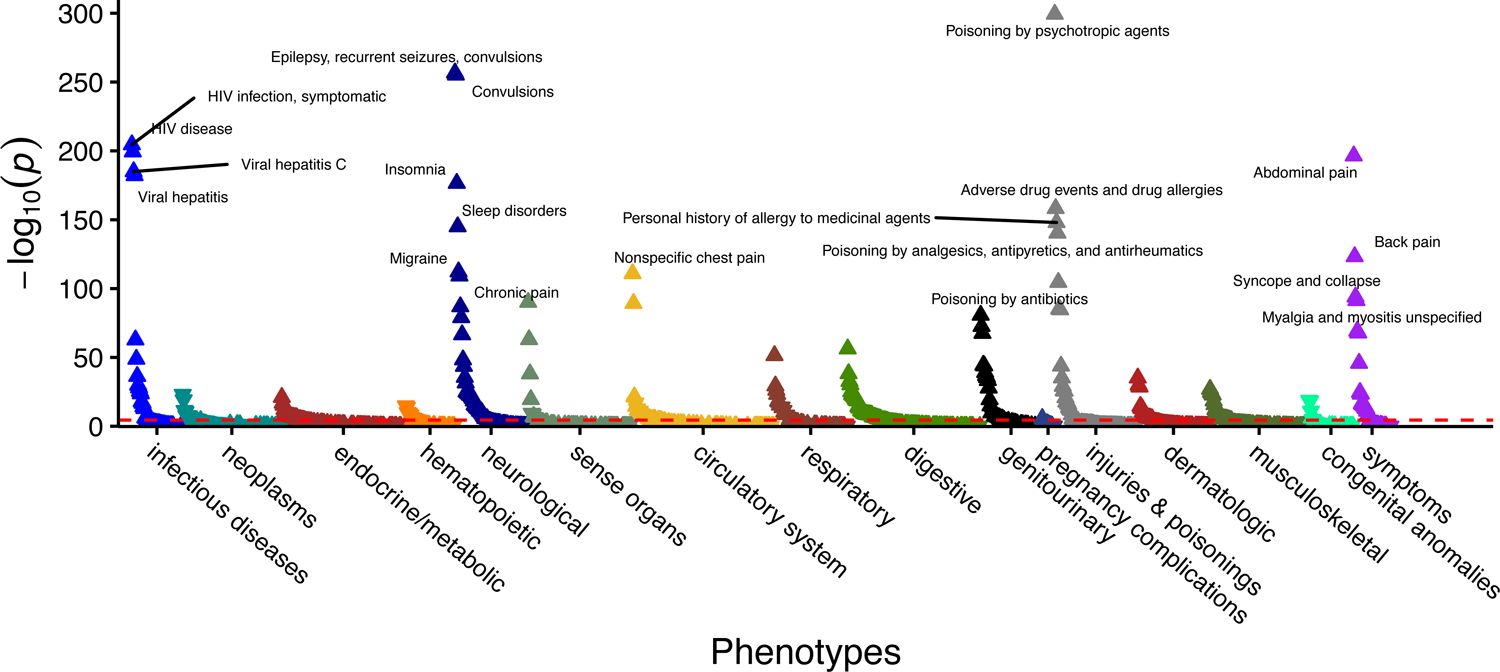
Non-psychiatric associations with sexual assault. PheWAS plot from sex-combined analysis displaying –log10(p-value) for all non-psychiatric phecodes analyzed. Dashed red line indicates significance threshold (Bonferroni-corrected p<0.05).

Survivors of sexual assault frequently suffer from disorders involving functional and somatic symptoms including chronic pelvic pain and chronic gastrointestinal symptoms^4^. In our analysis, the broad “somatoform disorder” phecode is strongly associated with sexual assault (OR=53.84, 95% CI=46.35, 62.55). We additionally note multiple clinical associations related to functional and somatic symptom disorders, including abdominal pain (OR=2.18, 95% CI=2.07, 2.29), chronic (nonspecific) pain (OR=2.24, 95% CI=2.09, 2.41), unspecified myalgia and myositis (OR=2.31, 95% CI=2.13, 2.5), and cervicalgia (OR=1.75, 95% CI=1.62, 1.88). We also identify a range of urinary symptoms associated with sexual assault, consistent with prior studies linking sexual assault and genitourinary symptoms^16,17^, including dysuria (OR=2.11, 95% CI=1.95, 2.29), urinary frequency (OR=2.18, 95% CI=1.95, 2.43), urinary retention (OR=2.67, 95% CI=2.33, 3.06), urinary incontinence (OR=1.97, 95% CI=1.78, 2.19), and cystitis (OR=2.35, 95% CI=2.06, 2.68).

Several phenome-wide-significant phecodes identified in our analysis pertained to seizures and epilepsy, including the broad “epilepsy, recurrent seizures, convulsions” phecode and all phecodes classified under it. Given the previously identified relationship between sexual assault and the development of functional seizures, we investigated whether these associations were partially explained by patients experiencing functional seizures as classified by our previously published phenotyping algorithm^10^. Indeed, we observed a high level of overlap between seizure-related phecodes and functional seizure status (**Supplemental Table 5)**. Furthermore, when we included functional seizure status as a covariate in the logistic regression models, all associations between sexual assault and seizure-related phecodes were at least partially attenuated (**Supplemental Table 6**).

### Sex-differential phenotypic association analyses

Sex differences in prevalence, age of onset, and presentation of many neuropsychiatric disorders are well characterized in the literature^18,19^. We analyzed our cohort for sex-differential phenotypic associations to determine whether a history of sexual assault is modified by sex assigned at birth. First, we first performed sex-stratified PheWAS of sexual assault disclosure across all phecodes with sufficient case counts (**Supplemental Table 4**, “male_only” and “female_only” sheets). We identified 410 phecodes significantly associated with sexual assault in at least one of the analyses, 332 of which were also significant in the sex-combined analysis.

Many of the significant sex-stratified associations were specific to one sex. We identified three male-specific associations with sexual assault: viral hepatitis A (OR=7.93, 95% CI=3.2, 19.64), testicular hypofunction (OR=2.25, 95% CI=1.73, 2.93), and testicular dysfunction (OR=2.24, 95% CI=1.72, 2.91). The female-specific associations were diverse and included infectious diseases, genitourinary conditions, and psychiatric disorders. Focusing on the psychiatric associations, we note that sexual assault is strongly associated with the broad “mental disorders during/after pregnancy” phecode (OR=28.77, 95% CI=24.6, 33.66). We also observe a strong association between sexual assault and the female-predominant diagnosis of dissociative disorder (OR=113.77, 95% CI=86.15, 150.24), which recapitulates previous findings that the majority of patients with this disorder have a history of abuse or neglect^20-21^ (cited in ^22^).

Of the 410 significant sex-stratified associations, the 354 phecodes with sufficient case counts in both sexes were tested in an interaction analysis examining the moderating effect of sex on sexual assault comorbidities. Of these, 54 phecodes demonstrated significant differences between males and females (Bonferroni-adjusted α=1.41e-04, **Figure 3, Supplemental Table 4** [“sex_interaction” sheet]), 32 of which pertained to psychiatric conditions, including depression, anxiety disorders, schizophrenia, and PTSD. Interestingly, in all 32 sex-differential psychiatric associations with sexual assault, the effect sizes were significantly greater in males than in females.

**Figure 3:**
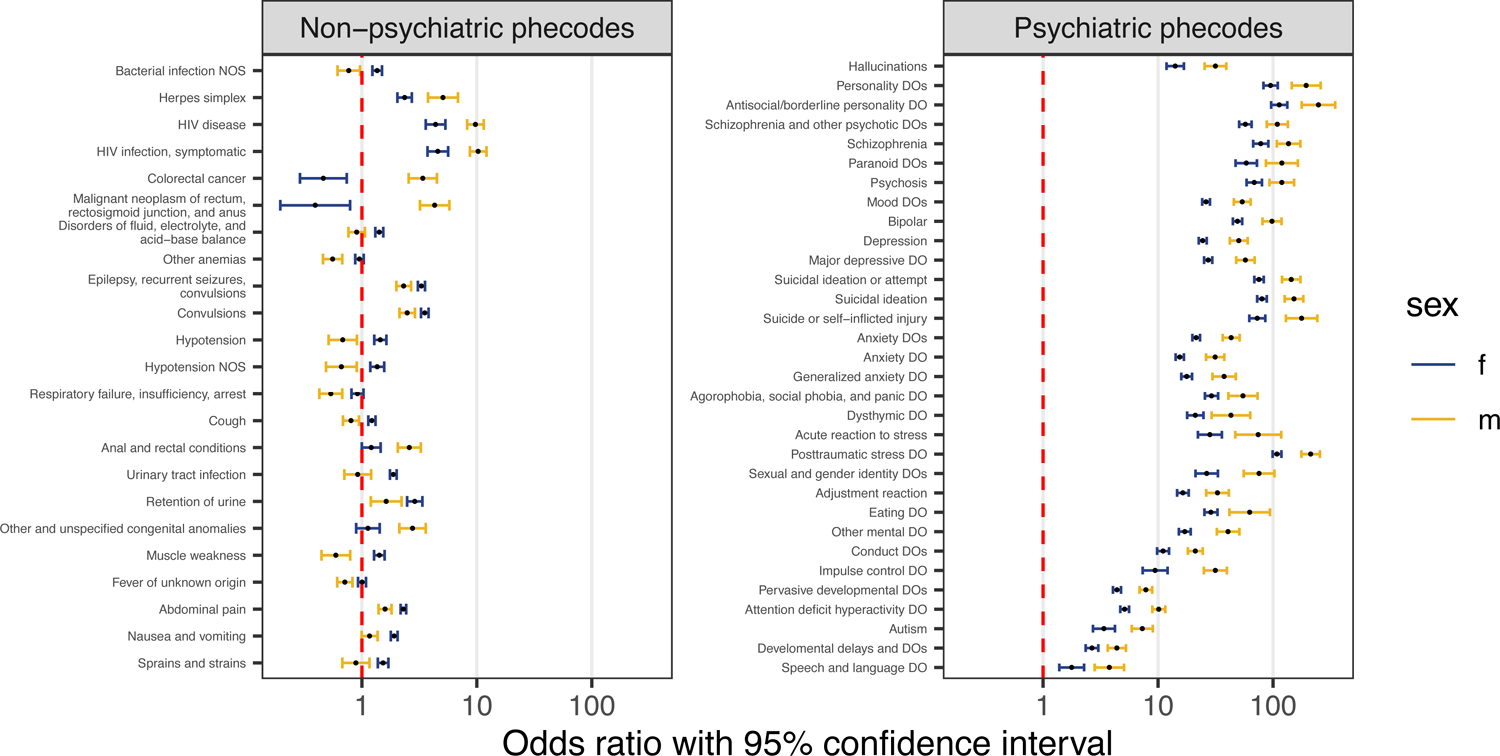
History of sexual assault increases odds of clinical phenotypes in a sex-differential manner. For each of the 55 phenotypes with significant sexual-assault-by-sex interaction effects, log10-scale odds ratio (OR) and 95% confidence interval is plotted separately by sex for non-psychiatric phecodes (left) and psychiatric phecodes (right). The ORs correspond to the sex-stratified associations between sexual assault and each phenotype. Associations are grouped by related phenotypes. Red dashed line indicates an OR of 1. (Note: DO=disorder).

Among the non-psychiatric associations, those stronger in females than in males included “sprains and strains,” gastrointestinal symptoms (“abdominal pain;” “nausea and vomiting”), urinary retention, urinary tract infection, and seizure-related phecodes (“epilepsy, recurrent seizures, convulsions;” “convulsions”). Non-psychiatric associations that were stronger in males included anal and rectal conditions, HIV-related phecodes, herpes simplex infection, and unspecified congenital anomalies. Several sex-differential associations with non-psychiatric phecodes exhibit opposite effects in men and women. For example, hypotension, unspecified bacterial infection, muscle weakness, and cough were observed more frequently in females affected by sexual assault than in females not affected, but the reverse was true for males. Conversely, colorectal cancer was observed more frequently in males with a sexual assault history than in those without, but the reverse was true for females.

### Benchmarking of keyword algorithm using ICD-code-based case-selection

To determine whether the associations identified by a keyword-based case-control definition are consistent with those identified by a simpler ICD-code-based approach, we repeated the PheWAS using ICD codes to classify patients as positive or negative for a history of sexual assault (see *Methods*). Of the 386 phenome-wide-significant sex-combined associations identified using the keyword-based algorithm, 289 were also nominally associated (p<.05) using the ICD-code-based algorithm. Furthermore, among phecodes identified as phenome-wide-significant by the keyword-based approach, log ORs from the two approaches were strongly correlated (Pearson R≥0.84, **Supplemental Figure 1, Supplemental Table 7**).

## Discussion

Epidemiological studies have established that individuals with a history of sexual assault have increased risk of developing both immediate and long-term physical and mental health conditions^2–4,7,8,22–24^. We aimed to replicate and extend these findings using a novel clinical informatics-based phenotyping approach in a large hospital setting. Our approach ascertained substantially more patients than did diagnostic codes alone, yielding improved power to study the medical phenomes of sexual assault survivors. This study not only confirmed the utility of our approach but also produced novel insights into the consequences of sexual assault for males and females.

Our phenotyping approach identified hundreds of clinical phenotypes significantly associated with sexual assault, including a preponderance of associations with psychiatric conditions such as schizophrenia, depression, PTSD, and suicidal behaviors. We observed several associations between sexual assault and phenotypes related to functional and somatic symptom disorders including gastrointestinal symptoms, chronic pain, and seizures. These results are consistent with findings from the few epidemiological studies examining the impact of sexual violence on the development of functional and somatic symptom disorders^4,8^. Nevertheless, they remain largely understudied and warrant further investigation.

We noted several phenotypes associated with sexual assault pertaining to toxic ingestions, such as poisoning by psychotropic agents and analgesics. Psychotropic agents including antidepressants, as well as commonly used analgesics such as paracetamol, are used frequently in suicide attempts^25,26^. Consistent with this, our analysis demonstrated a strong association between sexual assault and suicide and self-harm. Given the high overlap between toxic ingestions and documentation of suicidal behavior in our data (**Supplemental Table 8**), the associations between sexual assault and toxic ingestions are likely in part driven by suicidal behavior. Furthermore, when we conditioned on the “suicide or self-inflicted injury” phecode, these associations were diminished (**Supplemental Table 9**).

Finally, we addressed a gap in the sexual assault literature by leveraging our approach to study sex differences in risk factors and comorbidity patterns among survivors of sexual assault. We found that dissociative disorder, which occurs predominantly in women, is strongly associated with sexual assault in our cohort, recapitulating the observation that most women with the disorder have a history of abuse or neglect^20–22^. Our interaction analysis revealed that abdominal pain, nausea and vomiting, epilepsy, recurrent seizures, and convulsions are more commonly observed in female than male survivors of sexual assault. While we cannot conclusively assert that these conditions represent functional or somatic symptom disorders, prior work linking sexual assault to this class of disorders^4^ makes this interpretation plausible. Furthermore, we showed that the seizure-related associations are partially attenuated when conditioning on functional seizures case-control status, the phenotyping algorithm for which we recently published^10^.

Interestingly, most of the sex-differential psychiatric associations with sexual assault were stronger in men than in women. This finding should be interpreted in light of possible differential reporting and recall bias between males and females who have experienced sexual assault. For example, if males tend to report experiences of sexual assault less frequently than females, it is possible that the men who do disclose such a history in a medical setting are those who are experiencing especially severe health consequences as a result of their trauma. In this scenario, observed clinical associations with sexual assault would appear stronger in males than in females, which is what we observe for many sex-differential phenotypes in our analysis (in particular, psychiatric conditions).

## Limitations

The cross-sectional design of our study prohibits us from drawing conclusions about causal or temporal relationships between sexual assault and associated phenotypes. Some of the phenotypic associations we identified, including phecodes relating to congenital anomalies and neurodevelopmental disorders, are likely to have preceded sexual assault and could be interpreted as risk factors for experiencing sexual assault later in life. However, many of the identified associations are consistent with health sequelae of sexual assault identified in longitudinal case-control and cohort studies.

Secondly, our analysis is subject to the ascertainment bias inherent in EHR-based medical phenome studies. It is difficult to extrapolate our results, which are derived from a cohort of individuals seeking medical care, to the broader population. For example, survivors of sexual assault who frequently interact with the healthcare system may be more likely to receive a greater number of medical diagnoses than those who do not seek or have limited access to care. Nevertheless, it is critical that health care systems are prepared to anticipate the care needs of survivors of sexual assault to provide high quality primary preventative care. Moreover, we have attempted to account for this bias by including a measure of healthcare utilization as a covariate in our analyses.

Thirdly, the small number of cases identified by our keyword-based algorithm relative to sexual assault prevalence estimates^1^ suggests that, though it is an improvement over the use of ICD codes alone, our algorithm is still misclassifying many cases as controls and underestimating the prevalence of sexual assault in the VUMC patient population. This underestimation likely stems from ubiquitous underreporting of sexual assault in healthcare settings. For example, only approximately one third of women with injuries due to rape seek any type of medical care^27^. In the context of this misclassification, our analyses of clinical associations with sexual assault are likely to be overly conservative. While an overly conservative analysis introduces type II error, it does not reduce confidence in the positive clinical associations we have identified.

## Public health implications

Sexual assault is an urgent public health concern with both immediate and enduring physical and mental health consequences. Our study provides novel evidence supporting a strong relationship between sexual assault and psychiatric conditions, including functional and somatic symptom disorders. In our cohort, the associations between sexual assault history and psychiatric conditions are stronger in males than in females, which may reflect reporting bias and/or sex-differential biopsychological responses to trauma. Conversely, associations relating to gastrointestinal symptoms and seizures, which may represent manifestations of functional and somatic symptom disorders, were more commonly observed in female than male survivors of sexual assault.

Our analysis, in context of prior epidemiological studies, suggests that patients who have experienced sexual assault are at risk for developing wide-ranging medical and psychiatric comorbidities. Yet, sexual assault continues to be vastly underreported in healthcare settings. Further research is needed to understand the conditions under which patients feel safe in disclosing a history of sexual assault trauma. Furthermore, safe, compassionate, and effective protocols are needed for providers to routinely screen survivors for a variety of physical and mental health conditions, both immediately and longitudinally.

## Supporting information

Supplementary content

## Data Availability

All summary-level data produced in the present work are contained in the manuscript. De-identified VUMC electronic health record data analyzed in the present work are not available for public access.

## Figure and table legends

**Supplemental Table 1**. Criteria used in developing algorithm to identify patients with a history of sexual assault. Patients were identified using natural language processing of clinical notes using various inclusion phrases and exclusion phrases, shown here. The “Phase 1” case definition additionally included patients with matches to relevant ICD-9 and ICD-10 codes and yielded a positive predictive value (PPV) of 64% derived from 25 manually reviewed patient charts. The ICD codes used for the Phase 1 algorithm were all codes listed in Supplemental Table 2 except three codes (E960.1, Z04.41, Z04.42) which were identified later in the analysis process. False positives were used to refine the search terms, resulting in the final or “Phase 2” algorithm. The Phase 2 algorithm relies solely on natural language processing and does not take ICD codes into account. The PPV of the Phase 2 algorithm, based on manual review of 52 charts, was 90.4%.

**Supplemental Table 2** ICD-9 and ICD-10 codes used to identify sexual assault patients in the ICD-code-based algorithm. * indicates codes from the ICD-9 code set; all other codes listed are ICD-10.

**Supplemental Table 3** Demographics of patients classified as sexual assault cases or controls based only on relevant ICD codes. To assess case-control differences in demographic variables, two-sided t-tests were performed for continuous variables (record-median age, record-median BMI, log(mean records per day)), and chi-square tests were performed for binary variables (sex, white/non-Hispanic). Log(mean records per day) is a measure of the density of diagnostic codes across an individual’s health record.

**Supplemental Table 4** PheWAS summary statistics for sex-combined, male-only, female-only, and sexual-assault-by-sex interaction analyses. Here, the keyword-based sexual assault case definition was applied. In the sex_interaction tab, ORs, confidence intervals, and p-values derived from the sex-stratified analyses are re-displayed (OR.m, p.value.f, etc.). (OR=odds ratio, CI=confidence interval, “.f” = female-only analysis, “.m” = male-only analysis, “.int” = interaction analysis.)

**Supplemental Table 5** Contingency tables demonstrating co-incidence of seizure-related phecodes and functional seizure cases (functional seizure phenotyping algorithm described in ^10^). For a given phenotype, “NA” corresponds to individuals excluded from case-control analyses for that phenotype (see ^11^).

**Supplemental Table 6** Sex-combined associations between sexual assault and seizure-related phecodes after conditioning on functional seizures case-control status (functional seizure phenotyping algorithm described in ^10^). Here, the keyword-based sexual assault case definition was applied. (OR=odds ratio, CI=confidence interval.)

**Supplemental Figure 1**. Correlation scatterplots of log ORs from PheWAS on keyword-based (y-axis) and ICD-code-based (x-axis) case definitions. Log ORs are plotted for keyword-based associations meeting Bonferroni significance threshold. Results are reported separately for sex-combined, male-only, and female-only analyses.

**Supplemental Table 7** PheWAS summary statistics for sex-combined, male-only, and female-only analyses. Here, the ICD-code-based sexual assault case definition was applied. (OR=odds ratio, CI=confidence interval, “.f” = female-only analysis, “.m” = male-only analysis.)

**Supplemental Table 8** Contingency tables demonstrating co-incidence of toxic ingestions and suicidal behavior. For a given phenotype, “NA” corresponds to individuals excluded from case-control analyses for that phenotype (see ^11^).

**Supplemental Table 9** Sex-combined associations between sexual assault and toxic ingestions after conditioning on the “suicide or self-inflicted injury” phecode. Here, the keyword-based sexual assault case definition was applied. (OR=odds ratio, CI=confidence interval.)

## Acknowledgements

Research reported in this publication was supported by NIGMS of the National Institutes of Health under award number T32GM007347 and Grant No. R01 HG011405, partially funded by the Office of Research on Women’s Health, Office of the Director, NIH and the National Human Genome Research Institute. Its contents are solely the responsibility of the authors and do not necessarily represent the official views of the Office of Research on Women’s Health, the National Human Genome Research Institute, or the NIH.

