## Supplementary content for "Health conditions associated with sexual assault in a large hospital population"

Supplemental Table 1

| | Include | Exclude |
|---|---|---|
| **Phase 1** | molest(ed OR ation) | denie(s OR d) history of sexual (assault OR abuse) |
| | ‘ rape ’ | no history of sexual (assault OR abuse) |
| | sexual (harassment OR assault OR abuse) | sexual (abuse OR assault):(negative OR none) |
| | sexually (assaulted OR abused) | sexual (abuse OR assault): (negative OR none) |
| **Phase 2** | (history OR hx OR h/o) of sexual abuse | no (history OR hx OR h/o) of sexual abuse |
| | (history OR hx OR h/o) of sexual assault | denies (history OR hx OR h/o) of sexual abuse |
| | sexual (OR sexually) abuse(d) by | no (history OR hx OR h/o) of sexual assault |
| | (reports OR reported) a rape | denies (history OR hx OR h/o) of sexual assault |
| | (her OR his) rape | |
| | was raped | |
| | sexually abused (him OR her) | |
| | secondary to rape (OR sexual abuse OR sexual assault) | |

### Supplemental Table 2

| ICD Code | Description | ICD Code | Description |
|---|---|---|---|
| **E960.1*** | Rape | **T76.21XD** | Adult sexual abuse, suspected, subsequent encounter |
| **V71.5*** | Observation following alleged rape or seduction | **T76.21XS** | Adult sexual abuse, suspected, sequela |
| **995.83*** | Adult sexual abuse | **T76.22XA** | Child sexual abuse, suspected, initial encounter |
| **995.53*** | Child sexual abuse | **T76.22XD** | Child sexual abuse, suspected, subsequent encounter |
| **Z04.41** | Encounter for examination and observation following alleged adult rape | **T76.22XS** | Child sexual abuse, suspected, sequela |
| **Z04.42** | Encounter for examination and observation following alleged child rape | **Z62.810** | Personal history of physical and sexual abuse in childhood |
| **T74.22XA** | Child sexual abuse, confirmed, initial encounter | **Z91.410** | Personal history of adult physical and sexual abuse |
| **T74.22XD** | Child sexual abuse, confirmed, subsequent encounter | **O9A.411** | Sexual abuse complicating pregnancy, first trimester |
| **T74.22XS** | Child sexual abuse, confirmed, sequela | **O9A.412** | Sexual abuse complicating pregnancy, second trimester |
| **T74.21XA** | Adult sexual abuse, confirmed, initial encounter | **O9A.413** | Sexual abuse complicating pregnancy, third trimester |
| **T74.21XD** | Adult sexual abuse, confirmed, subsequent encounter | **O9A.419** | Sexual abuse complicating pregnancy, unspecified trimester |
| **T74.21XS** | Adult sexual abuse, confirmed, sequela | **O9A.42** | Sexual abuse complicating childbirth |
| **T76.21XA** | Adult sexual abuse, suspected, initial encounter | **O9A.43** | Sexual abuse complicating puerperium |

Supplemental Table 3

| | Case (N=4422) | Control (N=828763) | Test statistic | P-value |
|---|---|---|---|---|
| **Sex** | | | | |
| Female | 3318 (75.0%) | 471909 (56.9%) | $\chi^2$=586.83 | <0.001 |
| Male | 1104 (25.0%) | 356854 (43.1%) | | |
| **Record-median age** | | | | |
| Median [Min, Max] | 23.0 [0, 85.0] | 37.0 [0, 89.0] | t=-42.68 | <0.001 |
| **Record-median BMI** | | | | |
| Median [Min, Max] | 25.1 [10.5, 59.4] | 25.8 [10.0, 60.0] | t=0.37 | 0.709 |
| **White and non-Hispanic** | | | | |
| No | 1366 (30.9%) | 208733 (25.2%) | $\chi^2$=75.61 | <0.001 |
| Yes | 3056 (69.1%) | 620030 (74.8%) | | |
| **Log(mean records per day)** | | | | |
| Median [Min, Max] | -3.28 [-7.25, 0.292] | -4.08 [-7.68, 1.37] | t=47.18 | <0.001 |

### Supplemental Table 5

| | Functional seizures | | | |
|---|---|---|---|---|
| | **All** | **TRUE** | **FALSE** | **NA** |
| Total | 630152 | 1630 | 595943 | 32579 |
| **Epilepsy, recurrent seizures, convulsions** | | | | |
| TRUE | 29207 | 1474 | 6532 | 21201 |
| FALSE | 517777 | 3 | 517440 | 334 |
| NA | 83168 | 153 | 71971 | 11044 |
| **Convulsions** | | | | |
| TRUE | 24022 | 1455 | 4480 | 18087 |
| FALSE | 517777 | 3 | 517440 | 334 |
| NA | 88353 | 172 | 74023 | 14158 |
| **Epilepsy** | | | | |
| TRUE | 13106 | 235 | 1961 | 10910 |
| FALSE | 517777 | 3 | 517440 | 334 |
| NA | 99269 | 1392 | 76542 | 21335 |
| **Generalized convulsive epilepsy** | | | | |
| TRUE | 5921 | 26 | 451 | 5444 |
| FALSE | 517777 | 3 | 517440 | 334 |
| NA | 106454 | 1601 | 78052 | 26801 |
| **Partial epilepsy** | | | | |
| TRUE | 8380 | 130 | 840 | 7410 |
| FALSE | 517777 | 3 | 517440 | 334 |
| NA | 103995 | 1497 | 77663 | 24835 |

### Supplemental Table 6

| Phenotype | Description | OR | CI.lower | CI.upper | P-value |
|---|---|---|---|---|---|
| 345 | Epilepsy, recurrent seizures, convulsions | 2.73 | 2.38 | 3.13 | 1.35e-46 |
| 345.3 | Convulsions | 2.78 | 2.37 | 3.27 | 2.58e-35 |
| 345.1 | Epilepsy | 2.08 | 1.57 | 2.76 | 3.38e-07 |
| 345.11 | Generalized convulsive epilepsy | 1.59 | 0.85 | 2.97 | 1.49e-01 |
| 345.12 | Partial epilepsy | 2.02 | 1.29 | 3.14 | 2.00e-03 |

Supplemental Figure 1

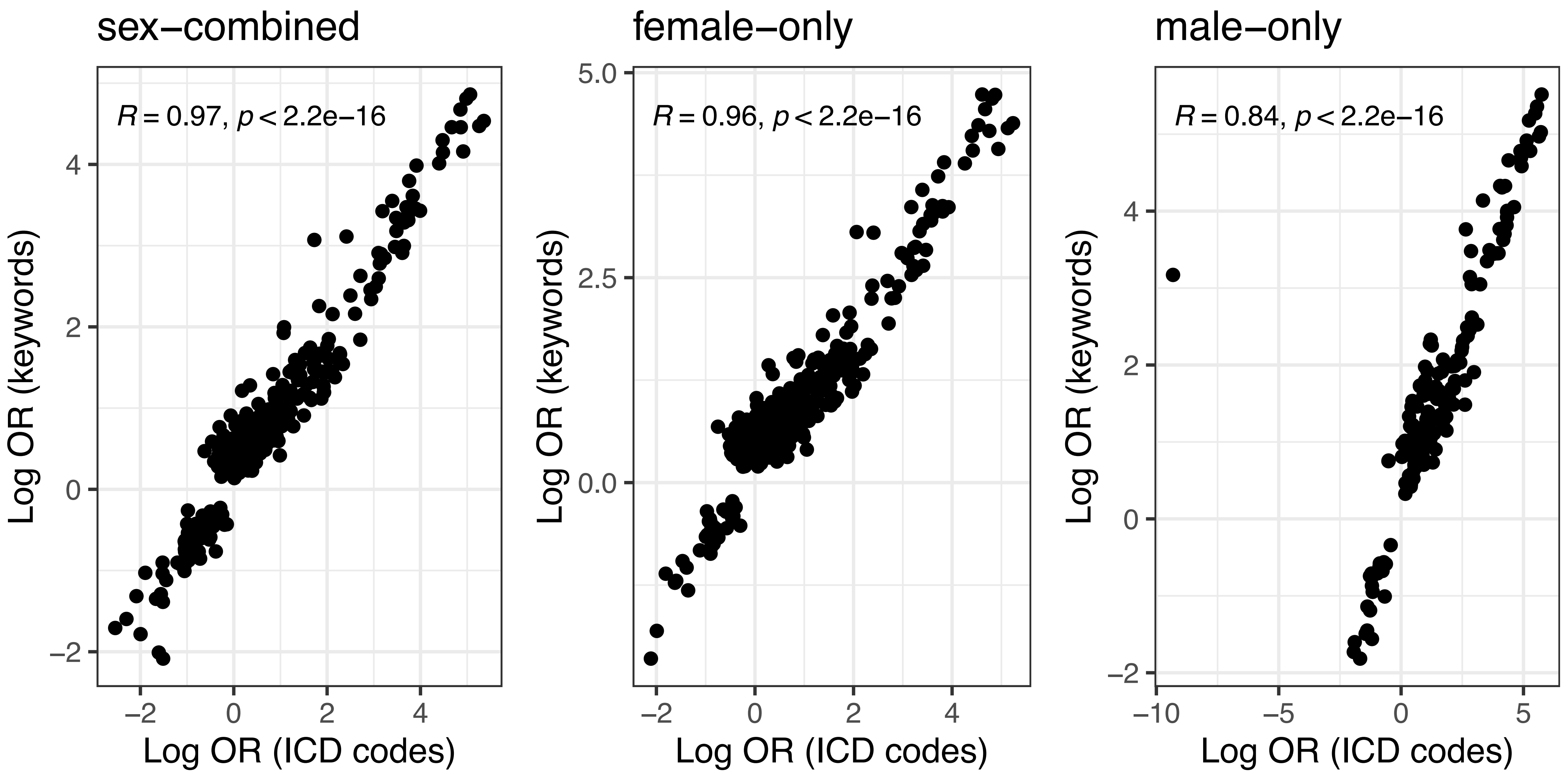

### Supplemental Table 8

| | Suicide or self-inflicted injury | | | |
|---|---|---|---|---|
| | **All** | **TRUE** | **FALSE** | **NA** |
| Total | 833185 | 1615 | 561158 | 270412 |
| **Poisoning by psychotropic agents** | | | | |
| TRUE | 1161 | 580 | 80 | 501 |
| FALSE | 709517 | 183 | 510264 | 199070 |
| NA | 122507 | 852 | 50814 | 70841 |
| **Poisoning by analgesics, antipyretics, and antirheumatics** | | | | |
| TRUE | 4959 | 389 | 1034 | 3536 |
| FALSE | 709517 | 183 | 510264 | 199070 |
| NA | 118709 | 1043 | 49860 | 67806 |
| **Poisoning by antibiotics** | | | | |
| TRUE | 14592 | 100 | 4217 | 10275 |
| FALSE | 709517 | 183 | 510264 | 199070 |
| NA | 109076 | 1332 | 46677 | 61067 |

| | Suicide or self-inflicted injury | | | |
|---|---|---|---|---|
| | **All** | **TRUE** | **FALSE** | **NA** |
| Total | 833185 | 1615 | 561158 | 270412 |
| **Poisoning/allergy of sulfonamides** | | | | |
| TRUE | 5663 | 28 | 1387 | 4248 |
| FALSE | 709517 | 183 | 510264 | 199070 |
| NA | 118005 | 1404 | 49507 | 67094 |
| **Poisoning by other anti-infectives** | | | | |
| TRUE | 573 | 7 | 114 | 452 |
| FALSE | 709517 | 183 | 510264 | 199070 |
| NA | 123095 | 1425 | 50780 | 70890 |
| **Poisoning by hormones and synthetic substitutes** | | | | |
| TRUE | 912 | 29 | 269 | 614 |
| FALSE | 709517 | 183 | 510264 | 199070 |
| NA | 122756 | 1403 | 50625 | 70728 |

Supplemental Table 9

| Phenotype | Description | OR | CI.lower | CI.upper | P-value |
|---|---|---|---|---|---|
| 969 | Poisoning by psychotropic agents | 2.03 | 1.22 | 3.36 | 6.09e-03 |
| 965 | Poisoning by analgesics, antipyretics, and antirheumatics | 1.70 | 1.01 | 2.84 | 4.51e-02 |
| 960 | Poisoning by antibiotics | 2.62 | 1.78 | 3.84 | 9.72e-07 |
| 961.1 | Poisoning/allergy of sulfonamides | 2.88 | 1.50 | 5.51 | 1.44e-03 |
| 961 | Poisoning by other anti-infectives | 3.47 | 0.81 | 14.79 | 9.28e-02 |
| 962 | Poisoning by hormones and synthetic substitutes | 3.24 | 1.37 | 7.68 | 7.60e-03 |
